# Evaluating the Potential Clinical and Economic Impact of Clesrovimab for Infants in England and Wales

**DOI:** 10.64898/2025.12.03.25341225

**Authors:** Klodeta Kura, Colleen Burgess, Ian Matthews, Salisu Garba, Dawei Wang, John Lang, Dionysios Ntais, Yao-Hsuan Chen

**Affiliations:** HEDS Vaccines, Merck Sharp & Dohme (UK) Ltd., London EC2M 6UR, UK; HEDS Vaccines, Merck Sharp & Dohme LLC, West Point, PA 19486, USA; Value, Access, and Devolved Nations (VAD), London, UK., Merck Sharp & Dohme (UK) Ltd., London EC2M 6UR, UK; Merck Canada Inc., Kirkland, QC, Canada

## Abstract

**Background:** Respiratory syncytial virus (RSV) infects nearly all children under two years, causing mild lower respiratory tract infections (LRTIs) although serious disease, mortality and long-term recurrent wheezing can also occur. This study evaluates the health and economic impact of various RSV prevention strategies (monoclonal antibodies [mAb] such as clesrovimab, nirsevimab, palivizumab [high risk infants only] or maternal vaccination) in infants in England and Wales.

**Methods:** A previously published decision tree model was adapted to estimate RSV LRTIs and associated costs over one year among a cohort of infants entering their first RSV season in England and Wales. Clinical trial data were used for efficacy inputs alongside country-specific epidemiology and costs to assess the impact of year-round and seasonal mAb programs (without and with catch-up [for infants born out of season]) alongside maternal vaccination. Deterministic sensitivity analyses of incremental treatment costs and QALYs were performed.

**Results:** Seasonal administration of clesrovimab with and without catch-up led to fewer RSV-associated LRTI cases and lower treatment costs compared to other RSV intervention strategies.

Without catch-up, clesrovimab reduced RSV-LRTI cases by 18%-27% versus palivizumab, 3%-4% versus nirsevimab and 2%-13% versus maternal vaccine.

With catch-up, RSV-LRTI outcomes reductions by clesrovimab were greater: 60%-70% versus palivizumab, 14%-20% versus nirsevimab, and 43%-54% versus maternal vaccination. It also reduced treatment costs by £37.0 million compared to palivizumab, £4.0 million compared to nirsevimab, and £26.0 million compared to maternal vaccination. Coverage and efficacies were the most influential parameters.

**Conclusions:** Clesrovimab is projected to substantially reduce RSV-associated clinical outcomes and costs compared to both standard of care and potential new RSV prevention measures in England and Wales.

## Introduction

Human respiratory syncytial virus (RSV) is a seasonal virus with peak activity in the UK occurring between October and February, that infects nearly all children within the first two years of life [1-3]. Severe RSV disease can lead to higher risk of acquiring asthma or recurrent wheezing later in life [5]. RSV infection is a substantial contributor to the hospitalization of children in Europe and is one of the leading causes of death in infants worldwide [6]. An estimated 33.1 million episodes of LRTIs, 3.2 million hospitalizations, and 59,600 in-hospital deaths due to RSV occurred in children < 5 years of age globally in 2015 alone [7].

In the UK, between 2007-2017 there were approximately 118,363 annual hospital admissions with respiratory infections in children aged <2 years old, of which approximately 32,485 (27.4%) were attributable to RSV [8].

For many years the recombinant monoclonal antibody (mAb) palivizumab (Abbott Labs Ltd initially, now Astra Zeneca) was the standard of care for protection of high-risk children against RSV in the UK. However, up to five monthly doses of palivizumab are required through the duration of the RSV season, at high cost per dose as protection is short-lived. These factors, alongside the National Health Service (NHS) capacity constraints that multiple injections can cause at the peak of the virus season, can lead to increased exposure to nosocomial infections and/or transmission of external pathogens within the hospital.

A universal maternal RSV immunization programme with Abrysvo® (Pfizer) for >28 weeks of pregnancy was introduced in the UK in 2024 [9]. Recently, the UK NHS also announced the introduction of the mAb nirsevimab (Sanofi) for premature infants (<32 weeks) with the aim of replacing palivizumab, noting the higher efficacy, longer protection against RSV, and the need for a single infusion [10]. Nirsevimab will also be used in at-risk infants born pre-term and/or suffering from respiratory or heart disease or immunodeficiencies [1, 11, 12]. Single-dose prophylaxis of infants with nirsevimab and single-dose maternal vaccination (MV) during pregnancy are both recommended in the UK’s Green Book; both interventions impart passive immunity to full-term infants, thereby protecting them from RSV disease during the first few months following birth [12, 13]. In addition, a single-dose long-acting mAb (clesrovimab) (developed by Merck Sharp & Dohme) has been approved in the US and is currently under review in other countries [14].

The aim of this study was to expand the static cohort model adaptation of Lang *et al* to evaluate the public health impact of clesrovimab and other RSV interventions in England and Wales [19]. Model parameters were updated from the latest published literature and cost components were added for RSV treatment.

## Methods

A previously developed static cohort model for RSV disease prevention was adapted for a single representative birth cohort from England and Wales [18, 19] over a 12-month time horizon. All births registered across these two countries were included in the model, partitioned by GA and high-risk sub-groups [20]. The model stratified the birth cohort by gestational and high-risk condition: high-risk infants (infants born <34 weeks gestational age [wGA] and with either CHD or chronic lung disease [CLD], 0.20%), extremely preterm infants (<29 wGA, 0.44%), very preterm infants (29-31 wGA, 0.59%),moderately preterm infants (32-34 wGA, 0.94%), mildly preterm infants (35-36 wGA, 5.42%), and full term infants (≥37 wGA, 92.42%) [20, 21], see Table 1 and Section 1 of Supplemental Materials for details. Birth cohorts were also stratified by birth month.

**Table 1.**
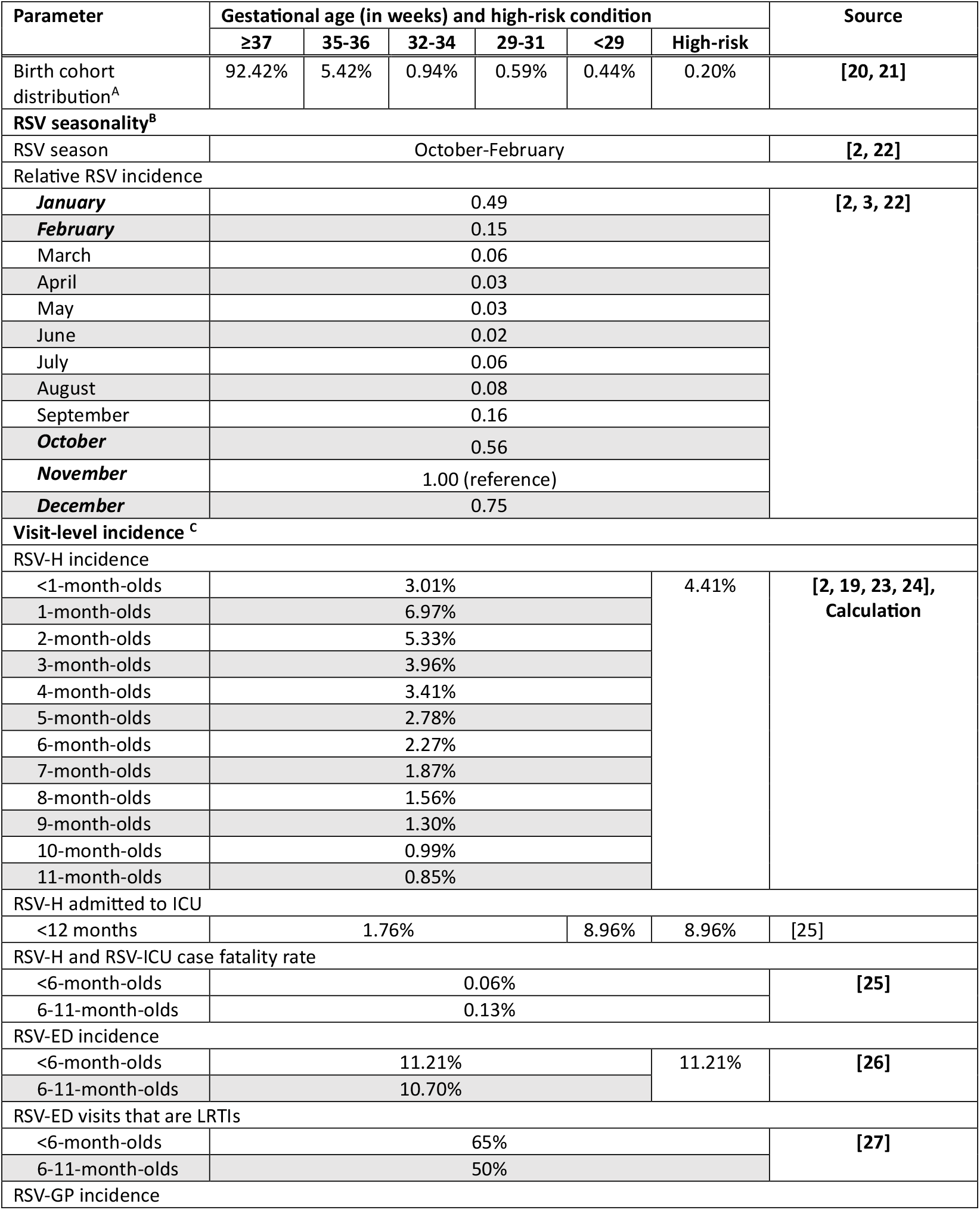
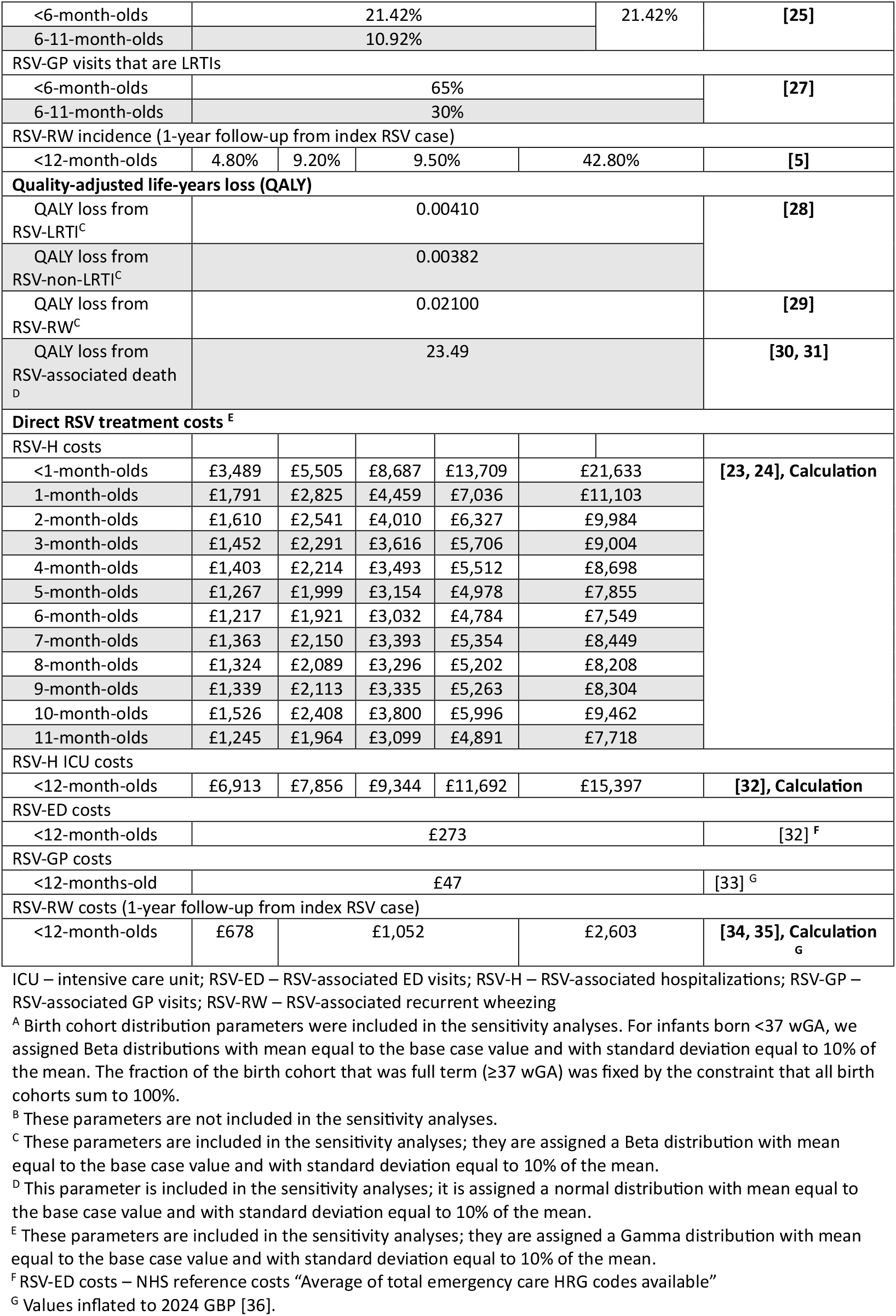
Model input parameters stratified by gestational age. All costs are shown as 2024 GBP.

### Model structure

The structure of the cohort model is a decision tree (Figure 1) in which infants in the various intervention arms may be infected with RSV and require medical attention in a general practice (GP), Accident & Emergency department (ED), or hospital inpatient (H) setting; a proportion of the latter group may then proceed to admission to an intensive care unit (ICU) or may die. Infants who survive a medically attended RSV episode may develop RSV-associated recurrent wheezing (RW) in the future.

**Figure 1.**
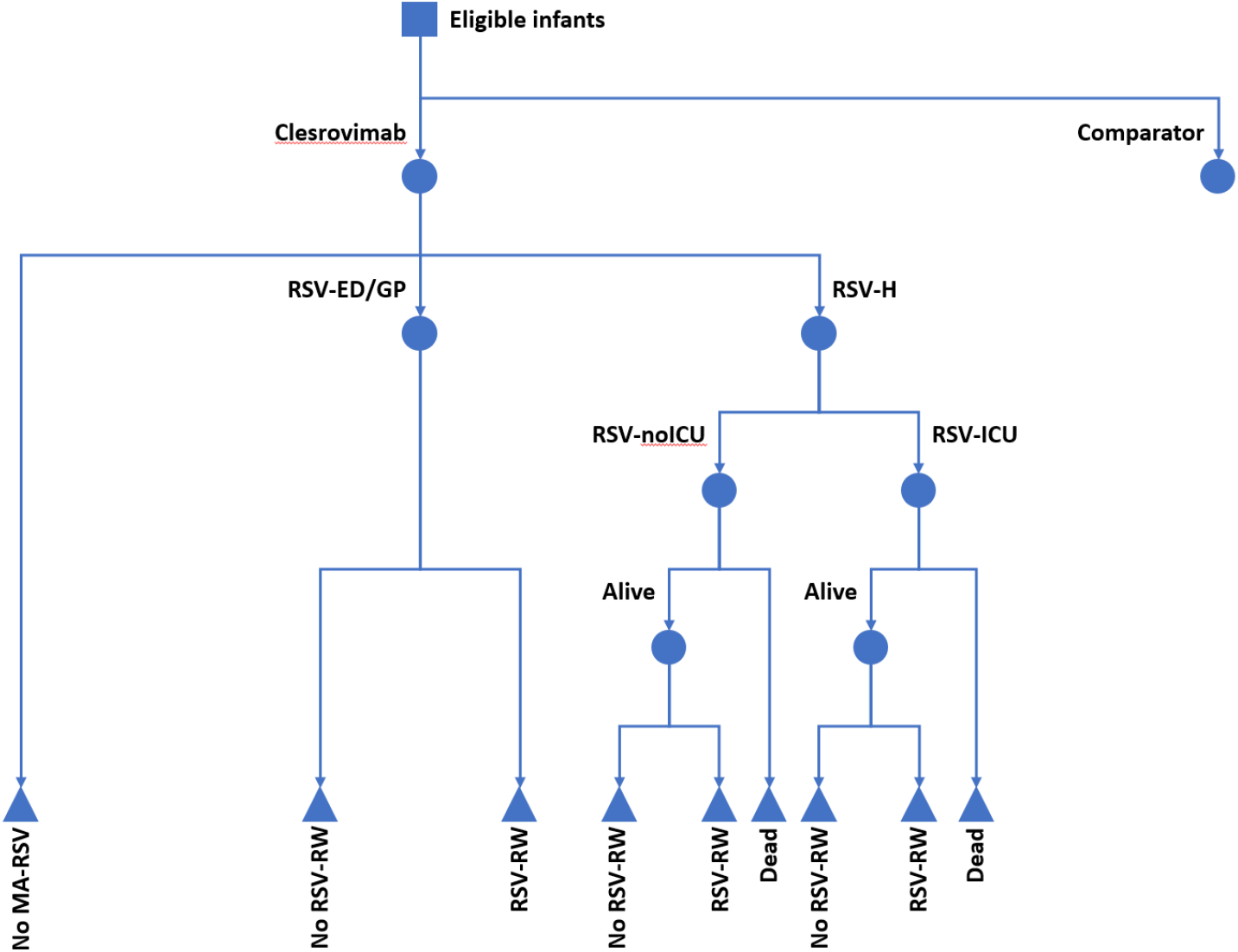
Decision tree structure for the RSV cohort model. (RVS-ED/GP: emergency department / general practitioner (outpatient) visit); RSV-H: hospitalization; ICU: intensive care unit; MA-RSV: medically attended RSV; RSV-RW: recurrent wheezing)

### Model inputs

The main inputs are described in Table 1, including (i) birth cohort, (ii) seasonality, (iii) incidence and cost data of RSV-H, RSV-ED, RSV-GP, and (iv) quality of life data. All costs were provided as 2024 GBP; refer to Supplement sections 6 and 7 for a list of unit costs from NHS ref cost database and /or other cost inputs inflated using the NHS cost inflation index [36].

### RSV intervention strategies

This model considers four different RSV interventions: palivizumab, clesrovimab, nirsevimab, and maternal vaccination (MV). The efficacy against medically attended lower respiratory infection (MALRI), uptake, and duration of protection of these interventions are summarized in Table 2.

**Table 2.** Intervention parameters.

| Prophylaxis | Uptake<sup>A</sup> | MALRI efficacy<sup>A</sup> | Duration of protection (months) |
| --- | --- | --- | --- |
| Palivizumab | 70% (55%,83%) [37] | 51% (37%,64%) [38] | 5 [38] |
| Clesrovimab | 90% (67%,100%) [19] | 87.2% (75.1%,93.4%) (versus nirsevimab)<br>75.1% (59.9%,84.5%) (versus maternal vaccine) [39, 40] | 6 [39, 40] |
| Nirsevimab | 90% (67%,100%) [19] | 79% (68.5%,86.1%) [41, 45, 46] | 5 [41] |
| Maternal vaccination | 54.3% (44%,67%) [42] | 51.3% (29.4%,66.8%) [43] | 6 [43] |

<sup>A</sup> Values in parentheses represent lower and upper bounds for the one-way sensitivity analysis. MALRI – medically attended lower respiratory infections.

### Palivizumab

Under prophylaxis with palivizumab, infants were eligible if they were born <34 wGA and had either congenital heart disease (CHD) or chronic lung disease (CLD). Palivizumab was assumed to be administered once per month for each month of the RSV season, up to a maximum of 5 administrations.

### Monoclonal antibody

Three mAb strategies were considered, applied to both clesrovimab and nirsevimab: seasonal administration, seasonal administration with catch-up, and year-round administration. Under the seasonal strategy infants were eligible for prophylaxis if they were born during the RSV season; administration of clesrovimab or nirsevimab was assumed to occur once at birth. Under the seasonal strategy with catch-up infants were eligible for prophylaxis if they were born during the RSV season or if they were <7 months old at the start of the RSV season (if born outside of season); for infants born during the RSV season, prophylaxis was assumed to be administered once at birth; for infants born prior to the RSV season, administration of clesrovimab or nirsevimab was assumed to occur once at the start of the RSV season. Under the year-round strategy all infants were eligible for prophylaxis, which was assumed to be administered once at birth, regardless of the RSV season. In the base case, we assumed that the duration of protection from mAb was equal to the duration of the randomized control trial (6 months for clesrovimab and 5 months for nirsevimab) [39, 40, 41].

Nirsevimab’s efficacy estimate against all MALRI is derived from pooled data from phase 2b and phase 3 clinical trials (79% for <6-months post-dose) [41, 45, 46].

Evaluating the impact of clesrovimab versus a specific comparator requires proper alignment between the endpoints used in the respective randomized control trials (RCTs). The endpoint that most closely aligns with the RCT endpoint used in the study for nirsevimab, is MALRI requiring ≥2 indicators of LRI/severity. Therefore, in all scenarios where clesrovimab was compared to nirsevimab, a constant efficacy of 87.2% for all gestational ages and for all MALRI outcomes (e.g., RSV-noICU, RSV-ICU, RSV-ED, and RSV-GP) for days 1-180 (equivalent to six months duration) post dose was applied.

In summary, efficacy was assumed to remain constant during the duration of protection observed in RCTs: 6 months for clesrovimab, and 5 months for nirsevimab. After these periods, efficacy was assumed to drop to zero. This approach likely underestimates protection from these interventions, since there is no biological reason for complete and instantaneous loss of protection following the initial 5–6-month observed follow up in the pivotal RCTs [44].

### Maternal vaccination (MV)

Maternal vaccine (MV) was administered to all eligible pregnant women regardless of timing with respect to the RSV season. Unlike palivizumab, clesrovimab, and nirsevimab the efficacy of the MV intervention was a function of gestational age at immunization and birth, since successful immunization depends on the transfer of maternal antibodies from mother to newborn. In short, infants born prematurely were likely to lack maternal antibodies since they were born prior to the successful transfer of maternal antibodies, see Section 4 of the Supplemental Materials. Aligning the endpoint for clesrovimab efficacy to that used for MV resulted in an efficacy estimate of 75.1 % (95% confidence interval: 59.9%-84.5%) for days 1-180 (i.e., for months 1-6) post dose [43].

### RSV seasonality

RSV exhibits seasonal trends and is more common in the UK during the winter months, with the season running from October through February, and peak incidence occurring in November [1-3, 22], see Section 2 of the Supplemental Materials.

### Model outcomes

The time horizon for this analysis was 12 months. All costs and effects were calculated over this time horizon, with the exception of RSV-associated deaths, for which QALY loss was calculated over the lifetime period (with a discount rate of 3.5%).

This analysis focused on RSV MALRI, excluding RSV non-LRTIs. The model outcomes included RSV-H (with and without ICU admission), RSV-ED, RSV-GP, and RSV-associated deaths. QALYs and health and economic outcomes averted between comparators were also estimated.

### Sensitivity Analysis

Deterministic sensitivity analyses (DSA) were performed for seasonal administration of clesrovimab versus seasonal administration of nirsevimab and year-round maternal vaccination for LRTIs only and excluding RW, to identify influential parameters with respect to incremental QALYs and incremental treatment costs. Each model parameter was assigned a distribution, see Table 1 and Table 2 for details. For the DSA, the upper and lower bounds for each parameter were set to the 2.5% and 97.5% quantiles of the assigned distribution.

## Results

### Epidemiological results

#### Seasonal administration

Under in-season palivizumab for at-risk infants, the model projected 18,811 RSV-related hospitalizations, 358 ICU admissions, more than 102,000 healthcare visits, 4,420 cases of recurrent wheezing, and 11 deaths — most occurring in full-term infants without CHD or CLD.

Seasonal administration of nirsevimab to all infants reduced these to 14,254 hospitalizations, 274 ICU admissions, 37,415 ED visits, 44,069 GP visits, 3,517 wheezing cases, and 9 deaths.

Clesrovimab, also given seasonally to all infants, further improved outcomes. Compared to palivizumab, it reduced hospitalizations and ICU admissions by 27.2% and 26.6%, respectively. Compared to nirsevimab, the reductions were 3.9% and 3.9%. ED and GP visits dropped by 18.2% and 26.9% versus palivizumab, and by 2.6% and 4.1% versus nirsevimab. Deaths were reduced by 27.0% and 3.9%, compared to palivizumab and nirsevimab, respectively (Table 3).

**Table 3.** Incremental change in RSV-associated clinical outcomes resulting from seasonal, seasonal plus catch-up, and year-round administration of clesrovimab to all infants, as compared with in-season administration of palivizumab to at-risk infants, seasonal and seasonal plus catch-up administration of nirsevimab to all infants, and year-round administration of maternal vaccine.

| Administration | Clinical outcome | Clesrovimab (87.2% MALRI efficacy) vs |  | Clesrovimab (75.1% MALRI efficacy) vs |
| --- | --- | --- | --- | --- |
|  |  | Palivizumab | Nirsevimab | Maternal vaccine* |
| Seasonal | RSV-H-noICU | -5,117 (-27.2%) | -560 (-3.9%) | -1,950 (-11.9%) |
|  | RSV-ED | -8,130 (-18.2%) | -954 (-2.6%) | -838 (-2.2%) |
|  | RSV-GP | -15,534 (-26.9%) | -1,824 (-4.1%) | -4,089 (-8.4%) |
|  | RSV-H-ICU | -95 (-26.6%) | -11 (-3.9%) | -41 (-12.9%) |
|  | RSV-RW | -1,025 (-23.2%) | -122 (-3.5%) | -335 (-8.6%) |
|  | RSV Deaths | -3 (-27.0%) | -0.3 (-3.9%) | -1 (-12.2%) |
| Seasonal plus catch-up | RSV-H-noICU | -13,047 (-69.4%) | -1,378 (-19.3%) | -8,780 (-53.6%) |
|  | RSV-ED | -26,472 (-59.4%) | -2,961 (-14.0%) | -16,635 (-43.3%) |
|  | RSV-GP | -40,106 (-69.4%) | -4,307 (-19.6%) | -25,251 (-52.1%) |
|  | RSV-H-ICU | -247 (-69.0%) | -26 (-19.2%) | -172 (-54.1%) |
|  | RSV-RW | -2,891 (-65.4%) | -317 (-17.2%) | -1,942 (-50.1%) |
|  | RSV Deaths | -8 (-69.3%) | -0.8 (-19.3%) | -5 (-53.7%) |
| Year-round | RSV-H-noICU | -10,381 (-55.2%) | -2,265 (-21.2%) | -6,309 (-38.5%) |
|  | RSV-ED | -19,758 (-44.3%) | -5,030 (-16.8%) | -10,756 (-28.0%) |
|  | RSV-GP | -35,265 (-61.0%) | -9,611 (-29.9%) | -20,553 (-42.4%) |
|  | RSV-H-ICU | -196 (-54.7%) | -43 (-21.1%) | -125 (-39.2%) |
|  | RSV-RW | -2,347 (-53.1%) | -602 (-22.5%) | -1,445 (-37.3%) |
|  | RSV Deaths | -6 (-55.1%) | -1.4 (-21.1%) | -4 (-38.7%) |

ICU – intensive care unit; RSV-ED – RSV-associated ED visits; RSV-H – RSV-associated hospitalizations; RSV-GP – RSV-associated GP visits; RSV-RW – RSV-associated recurrent wheezing
\* Administration of maternal vaccine is year-round.

Year-round maternal vaccination led to 16,369 hospitalizations, 318 ICU admissions, 86,947 healthcare visits, 3,879 wheezing cases, and 10 deaths. Compared to this, seasonal administration of clesrovimab reduced all outcomes by 2%-13% (Table 3).

#### Seasonal plus catch-up administration

Seasonal plus catch-up administration of clesrovimab was projected to reduce hospitalizations and ICU admissions by 69.4% and 69.0% versus palivizumab, and by 19.3% and 19.2% versus nirsevimab. ED visits dropped by 59.4% versus palivizumab and 14.0% versus nirsevimab; GP visits by 69.4% and 19.6%, respectively. Deaths were reduced by 69.3% versus palivizumab and 19.3% versus Nirsevimab (Table 3).

Compared to year-round maternal vaccination, clesrovimab reduced hospitalizations and ICU admissions by approximately 54%, ED and GP visits by approximately 43–52%, and deaths by 54% (Table 3).

#### Year-round administration

Year-round administration of nirsevimab to all infants led to 10,695 and 205 RSV-related hospitalizations and ICU admissions, 29,862 and 32,125 accident and emergency department and GP visits, 2,675 cases of recurrent wheezing, and 6 deaths.

Year-round clesrovimab reduced hospitalizations and ICU admissions by 55.2% and 54.7% versus in-season palivizumab, and by 21.2% and 21.1% versus year-round nirsevimab. ED and GP visits were reduced by 44.3% and 61.0% versus palivizumab, and by 16.8% and 29.9% versus nirsevimab. Deaths were reduced by 55.1% versus palivizumab and 21.1% versus nirsevimab (Table 3).

Compared to year-round maternal vaccination, clesrovimab reduced all outcomes by 28-42%, including a 39% reduction in deaths (Table 3).

Additional epidemiological results for maternal vaccination of mothers of infants born within season only are presented in Section 7 of the Supplemental Materials.

## Cost results

### Seasonal administration

In-season palivizumab use was projected to cost £54.8 million in direct treatment (2024 GBP), excluding RW, with 87.3% attributed to full-term infants without CHD or CLD. Treatment costs under seasonal administration of nirsevimab cost were estimated to be £40.7 million. Seasonal clesrovimab reduced treatment costs by £15.9 million versus palivizumab and £1.8 million versus nirsevimab, with the majority of savings seen in healthy infants (Table 4).

**Table 4.** Incremental change in RSV-associated costs resulting from seasonal, seasonal plus catch-up, and year-round administration of clesrovimab to all infants, as compared with in-season administration of palivizumab to at-risk infants, seasonal and seasonal plus catch-up administration of nirsevimab to all infants, and year-round administration of maternal vaccine. (2024 GBP) Table 1: Model input parameters stratified by gestational age. All costs are shown as 2024 GBP.

| Administration | Cost outcome | Clesrovimab (87.2% MALRI efficacy) vs |  | Clesrovimab (75.1% MALRI efficacy) vs |
| --- | --- | --- | --- | --- |
|  |  | Palivizumab | Nirsevimab | Maternal vaccine* |
| Seasonal | RSV-H-noICU | -£12,210,259 (-32.9%) | -£1,347,068 (-5.1%) | -£7,042,235 (-20.8%) |
|  | RSV-ED | -£2,219,189 (-18.2%) | -£260,519 (-2.6%) | -£228,671 (-2.2%) |
|  | RSV-GP | -£730,425 (-26.9%) | -£85,747 (-4.1%) | -£192,282 (-8.4%) |
|  | RSV-H-ICU | -£707,708 (-25.7%) | -£82,739 (-3.9%) | -£362,573 (-14.4%) |
|  | RSV-RW | NA | NA | NA |
|  | Total treatment cost | -£15,867,582 (-29.0%) | -£1,776,074 (-4.4%) | -£7,825,761 (-15.9%) |
|  | Doses administered | 222,429 | 0 | (104,954) |
|  | QALY loss averted | 191 (-24.9%) | 22 (-3.6%) | 57 (-8.7%) |
| Seasonal plus catch-up | RSV-H-noICU | -£25,977,135 (-70.0%) | -£2,762,440 (-19.9%) | -£18,898,799 (-55.9%) |
|  | RSV-ED | -£7,226,161 (-59.4%) | -£808,320 (-14.0%) | -£4,540,868 (-43.3%) |
|  | RSV-GP | -£1,885,772 (-69.4%) | -£202,510 (-19.6%) | -£1,187,311 (-52.1%) |
|  | RSV-H-ICU | -£1,891,020 (-68.6%) | -£204,913 (-19.2%) | -£1,381,686 (-54.9%) |
|  | RSV-RW | NA | NA | NA |
|  | Total treatment cost | -£36,980,089 (-67.5%) | -£3,978,183 (-18.3%) | -£26,008,665 (-53.0%) |
|  | Doses administered | 550,412 | 0 | 223,028 |
|  | QALY loss averted | 514 (-67.0%) | 55 (-17.9%) | 335 (-50.7%) |
| Year-round | RSV-H-noICU | -£21,512,848 (-57.9%) | -£4,183,064 (-21.1%) | -£14,661,660 (-43.4%) |
|  | RSV-ED | -£5,393,359 (-44.3%) | -£1,372,943 (-16.8%) | -£2,936,111 (-28.0%) |
|  | RSV-GP | -£1,658,157 (-61.0%) | -£451,891 (-29.9%) | -£966,394 (-42.4%) |
|  | RSV-H-ICU | -£1,490,075 (-54.1%) | -£335,440 (-20.9%) | -£1,011,607 (-40.2%) |
|  | RSV-RW | NA | NA | NA |
|  | Total treatment cost | -£30,054,439 (-54.9%) | -£6,343,338 (-20.4%) | -£19,575,773 (-39.9%) |
|  | Doses administered | 550,412 | 0 | 223,028 |
|  | QALY loss averted | 417 (-54.3%) | 102 (-22.5%) | 246 (-37.2%) |

ICU – intensive care unit; RSV-ED – RSV-associated ED visits; RSV-H – RSV-associated hospitalizations; RSV-GP – RSV-associated GP visits; RSV-RW – RSV-associated recurrent wheezing
\* Administration of maternal vaccine is year-round.

Year-round administration of maternal vaccine was projected to result in £49.1 million in treatment costs. In comparison, clesrovimab averted approximately £7.8 million in treatment costs (Table 4).

### Seasonal plus catch-up administration

Seasonal plus catch-up administration of nirsevimab was projected to lead to more than £21.8 million in treatment costs, excluding treatment of recurrent wheezing. In comparison, clesrovimab averted £37.0 million in treatment costs versus palivizumab and £4.0 million versus nirsevimab (Table 4).

Compared to year-round maternal vaccination, seasonal plus catch-up administration of clesrovimab saved £26.0 million in treatment costs (Table 4).

### Year-round administration

Year-round administration of nirsevimab was projected to lead to more than £31.0 million in treatment costs, excluding treatment of recurrent wheezing, with 86.1% of costs attributable to treatment of RSV in otherwise healthy infants. Year-round clesrovimab averted £30.1 million versus in-season palivizumab and £6.3 million versus nirsevimab, with most savings seen among healthy full-term infants. (Table 4).

Compared to year-round maternal vaccination, year-round administration of clesrovimab saved £19.6 million (Table 4).

Additional cost results for maternal vaccination of mothers of infants born within season only are presented in Section 7 of the Supplemental Materials.

### Sensitivity analyses

#### DSA for QALYs

A DSA was conducted comparing seasonal clesrovimab to seasonal nirsevimab (focusing on LRTIs only, excluding RW) to identify key factors affecting incremental QALYs. Compared to seasonal nirsevimab, QALYs gained with clesrovimab were most sensitive to its uptake and efficacy, as well as nirsevimab’s efficacy. Additionally, QALY loss per LRTI had a minor influence on outcomes (Figure 2).

**Figure 2.**
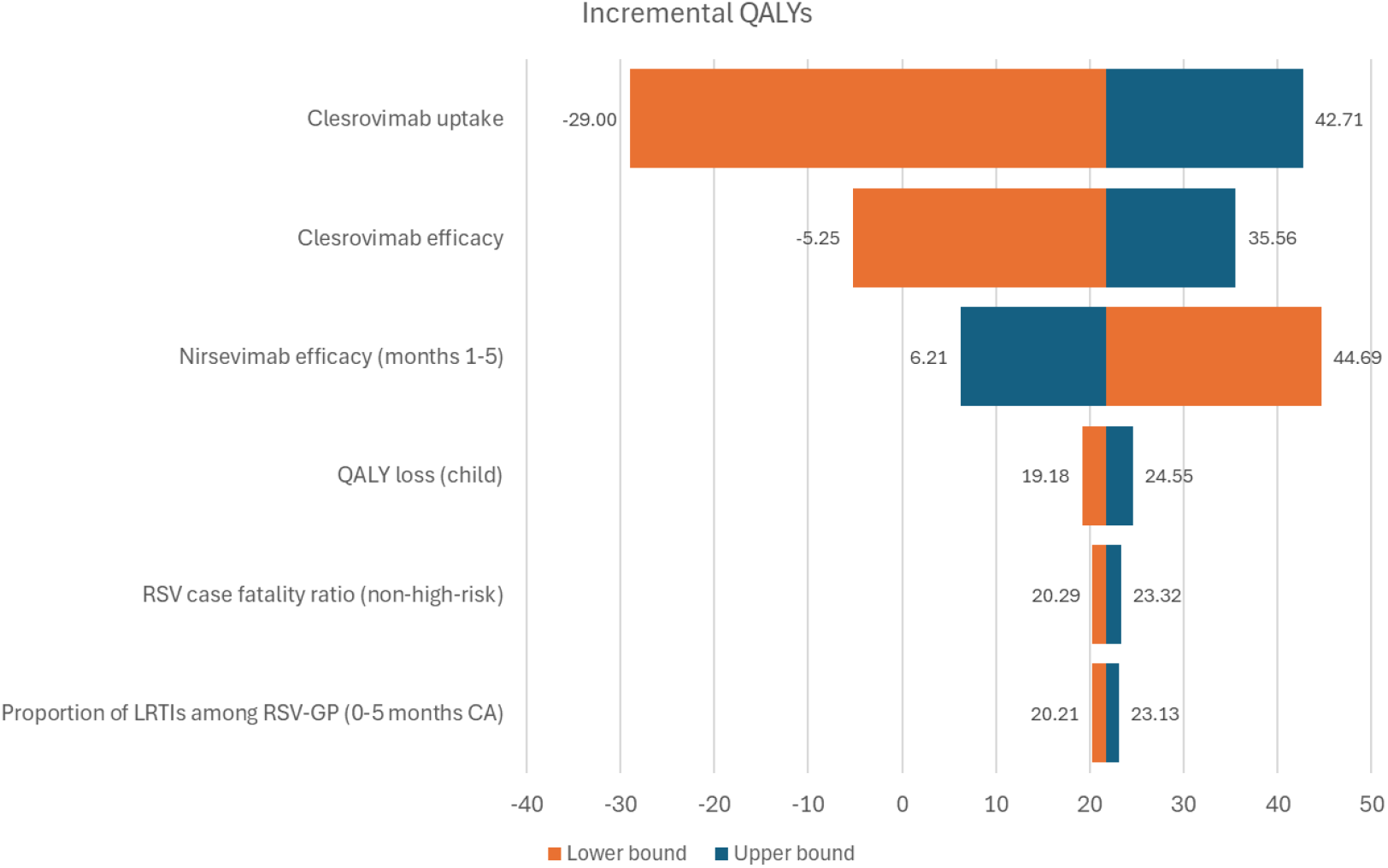
Incremental QALYs one-way sensitivity analysis for seasonal administration of clesrovimab versus seasonal administration of nirsevimab.

Additional DSA for QALYs results for maternal vaccination of mothers of infants born within season only are presented in Section 7 of the Supplemental Materials.

### DSA for costs

A DSA was conducted to assess incremental treatment costs of seasonal clesrovimab versus seasonal nirsevimab for LRTIs only (excluding RW). Compared to seasonal nirsevimab, results showed that treatment costs were most sensitive to clesrovimab uptake, with efficacy of both interventions also influencing the outcomes (Figure 3).

**Figure 3.**
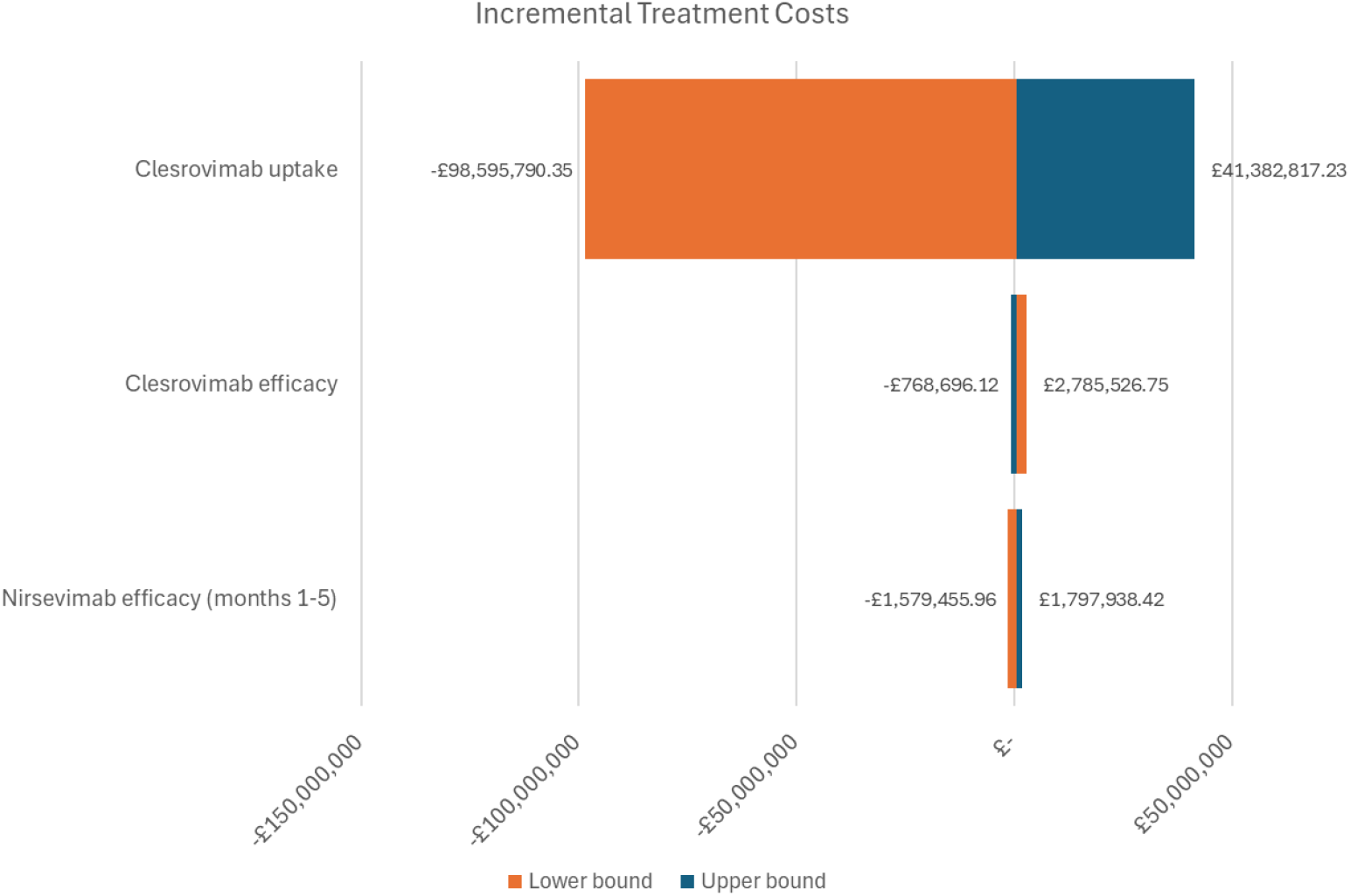
Incremental treatment costs one-way sensitivity analysis for seasonal administration of clesrovimab versus seasonal administration of nirsevimab. (2024 GBP)

Additional DSA for costs results for maternal vaccination of mothers of infants born within season only are presented in Section 7 of the Supplemental Materials.

## Discussion

This analysis adapted a previously published static model for the transmission and prophylaxis of RSV in infants to England and Wales, accounting for the simultaneous influences of chronological age and gestational age at birth, along with RSV seasonality, on an infant’s risk of RSV-associated outcomes and related costs [18, 19]. Model results suggest that, of the administration strategies evaluated, seasonal plus catch-up administration of clesrovimab led to the greatest proportional improvement in clinical outcomes and the lowest total treatment costs versus the comparators of in-season administration of palivizumab, seasonal plus catch-up administration of nirsevimab, and year-round maternal vaccination. In the UK RSV disease and treatment leads to significant clinical and economic burden in high-risk infants born pre-term and/or with CHD or CLD. However, there is also a high burden of RSV in low-risk full-term infants – this analysis projected that approximately 95% of RSV-related hospitalizations, 90% of ICU admissions, and 98% of outpatient and emergency department visits in children less than one year old in England and Wales would occur in low-risk infants, for all modeled treatments.

For nearly 25 years palivizumab was the standard of care for RSV prophylaxis in infants in England and Wales, targeting only those infants at high-risk of severe disease and leaving the majority of the infant population unprotected [1]. As newer alternatives were developed, mathematical models evaluating their clinical effectiveness and cost-effectiveness were published to help policy makers choose optimal intervention programs.

In 2020, Hodgson and colleagues developed a dynamic model of RSV transmission in the UK over a 10-year time horizon, including palivizumab for high-risk infants, and long-acting mAb and maternal vaccine as population-level intervention strategies, exploring the potential impacts of timing of implementation as well as the target population [2]. Their analysis projected that population-level administration of long-acting mAb or maternal vaccine could both be cost-effective replacements for palivizumab in the pediatric national immunization programme in England and Wales, depending on the cost per dose for each, and concluded that seasonal administration was the optimal approach for all interventions evaluated, preventing 28% of hospitalizations due to RSV [2]. The Hodgson and colleagues analysis was updated in 2022 to evaluate in greater detail the implementation and cost per dose of nirsevimab, again finding that seasonal administration would be the optimal prophylactic approach depending on intervention cost, with annual administration being cost-effective only if the duration of protection lasted nearly a year and if the cost of the annual programme was substantially lower than the seasonal one [15]. Following a thorough review of the analysis, the UK Joint Committee on Vaccination and Immunization determined the Hodgson model was conservative and should be updated with more recent inputs for costs and efficacy [12].The model was updated a third time in 2024 to include more recent efficacy trial data, finding that both maternal vaccine and long-acting mAb prophylaxes have the potential to significantly reduce RSV disease burden, with the latter outperforming the former under the assumption of higher coverage rate for long-acting mAb [16].

In contrast to these dynamic modeling approaches, Getaneh and colleagues developed a static model for RSV transmission in children under 5 years old in several European countries, to evaluate the cost-effectiveness of seasonal, seasonal with catch-up, and year-round administration of maternal vaccine and nirsevimab. The authors concluded that model results were “slightly more in favor of the seasonal [nirsevimab] program (October to April) with catch-up” than for the other modeled interventions, though the optimal programme varied depending upon the setting and the willingness-to-pay threshold [17]. The Getaneh analysis included different input assumptions from the Hodgson model, and reflected significant inter-country differences in cost-effectiveness results, along with high levels of uncertainty around hospitalization and quality-adjusted life-years (QALYs) lost per RSV episode. In 2022, Ektare and colleagues incorporated the simultaneous roles of chronological age and gestational age at birth, along with RSV seasonality, in an infant’s risk of RSV-associated outcomes and related costs into a static model for RSV transmission and prophylaxis in the United States [18]. Their analysis also found that full-term infants represented the highest proportion of RSV-associated clinical endpoints, and that long-acting mAb was the only intervention to adequately address disease impact in this population [18]. Following this, Lang *et al* adapted the model by Ektare and colleagues to the England and Wales setting, andperformed a comparative modelling analysis which compared the results of the static cohort model to those of the dynamic transmission model of Hodgson and colleagues [19]. The authors concluded that both static and dynamic models were appropriate for evaluating the impact of RSV prophylaxis.

The analysis described here adds to this body of literature in supporting the expansion of the RSV program within England and Wales to provide prophylaxis to all infants, with administration of the long-acting mAb clesrovimab leading to the greatest improvements in clinical and treatment costs.

This analysis is subject to several limitations. The model described here requires highly detailed input parameters stratified by GA, CA, birth month, and RSV seasonality. One of the primary limitations of this model is the lack of such stratification in the literature needed for inputs. For example, the incidence of outpatient and emergency department visits stratified by GA and CA were not available; thus, the model assumed these parameters were constant for all GA. As these data become available in the future, the model can be updated to reflect this increase in granularity. Next, the model does not differentiate between CHD and CLD as they pertain to RSV-associated risk, nor does it account for reinfection. Further, this analysis is not a cost-effectiveness analysis. Finally, the model does not account for the potential for herd immunity from MV as an additional benefit to both mothers and infants, which may lead to under- or over-estimation of medically attended RSV visits.

## Conclusions

Of the RSV interventions evaluated in this analysis, seasonal plus catch-up administration of clesrovimab was the most effective at reducing RSV-associated clinical endpoints in all infant age- and risk-groups. By simultaneously accounting for both GA and CA, along with seasonality of the RSV force of infection, our model provides a more accurate estimate of clinical and economic outcomes of RSV disease and allows for direct comparison between multiple intervention strategies to aid in the development of RSV prevention policies.

## Data Availability

All data produced in the present work are contained in the manuscript

## Acknowledgements

The authors would like to thank:

Kayla Engelbrechtl; VAD, MSD (UK) Ltd for her early contribution in data sourcing and curation for modeling needs.

Yoonyoung Choi; Outcomes Research, Merck & Co., Inc., Rahway, NJ, USA for her early contribution in study discussion.

Vicky Coles: VAD, MSD (UK) Ltd for her contribution in model literature comparisons.

## Authorship contribution statement

All authors attest they meet the ICMJE criteria for authorship.

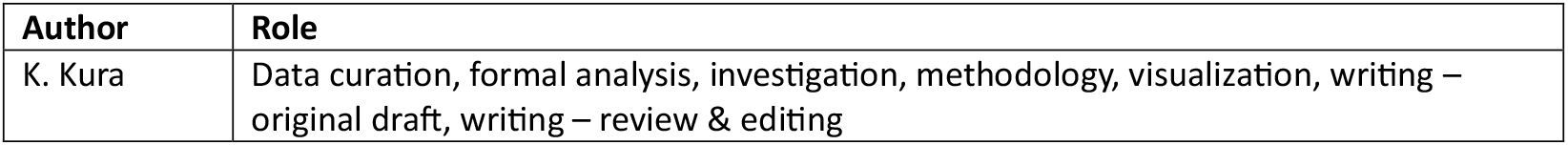
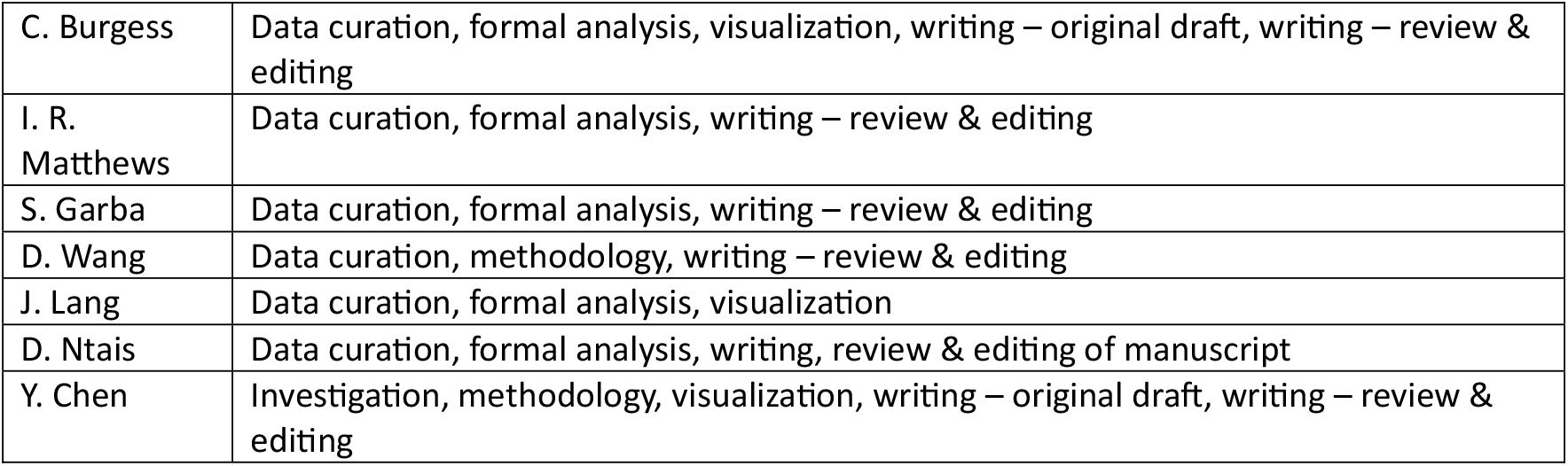

| Author | Role |
| --- | --- |
| K. Kura | Data curation, formal analysis, investigation, methodology, visualization, writing – original draft, writing – review & editing |
| C. Burgess | Data curation, formal analysis, visualization, writing – original draft, writing – review & editing |
| I. R. Matthews | Data curation, formal analysis, writing – review & editing |
| S. Garba | Data curation, formal analysis, writing – review & editing |
| D. Wang | Data curation, methodology, writing – review & editing |
| J. Lang | Data curation, formal analysis, visualization |
| D. Ntais | Data curation, formal analysis, writing, review & editing of manuscript |
| Y. Chen | Investigation, methodology, visualization, writing – original draft, writing – review & editing |

## Abbreviations

RSV: Respiratory syncytial virus
UK: United Kingdom
LRTI: Lower respiratory tract infection
CHD: Congenital heart disease
MV: Maternal vaccine
mAb: Monoclonal antibody
QALY: Quality adjusted life year
GA: Gestational age
CA: Chronological age
CLD: Chronic lung disease
ED: Accident and emergency department
GP: General practitioner
H: Hospitalization
ICU: Intensive care unit
RW: Recurrent wheezing
MA: Medically attended
MALRI: Medically attended lower respiratory infection
RCT: Randomized control trial
GBP: British pound sterling
WTP: Willingness to pay
DSA: Deterministic sensitivity analysis

## Notes

### Competing Interest Statement

KK, IM, DN and YHC are employees of MSD (UK) Limited, London, UK, a subsidiary of Merck & Co., Inc., Rahway, NJ, USA. JCL is an employee of Merck Canada Inc., a subsidiary of Merck & Co., Inc., Rahway, NJ, USA. SG, CB and DW are employees of Merck Sharp & Dohme LLC, a subsidiary of Merck & Co., Inc., Rahway, NJ, USA

### Funding Statement

This study was supported by Merck Sharp & Dohme Corp., a subsidiary of Merck & Co., Inc., Rahway, NJ, USA

