## Supplemental Materials for "Evaluating the Potential Clinical and Economic Impact of Clesrovimab for Infants in England and Wales"

Supplementary Materials

### 1. Demographic Model Inputs

Demographic model inputs are summarized in Appendix Table 1. Stratification by gestational age and high-risk medical condition (born at <34 weeks gestational age and with either congenital heart disease [CHD] or chronic lung disease of [CHD]) were previously estimated by Lang and colleagues [1] and are reproduced below.

*Appendix Table 1: Demographic model parameters*

| Parameter | Base case value | Distribution[A] | Reference |
|---|---|---|---|
| Total annual births | 624,729 | Normal | [2] |
| **Births by GA and high-risk condition** | | | |
| ≥37 wGA | 92.42% | N/A[B] | [2, 3], calculated below |
| 35-36 wGA | 5.42% | Beta | |
| 32-34 wGA | 0.94% | | |
| 29-31 wGA | 0.59% | | |
| <29 wGA | 0.44% | | |
| High-risk | 0.20% | | |

GA – gestational age; wGA – weeks gestational age

[A] Distributions are parameterized by assuming the mean is equal to the base case value and the standard deviation is equal to 10% of the mean.

[B] For sensitivity analyses the fraction of the birth cohort that is born ≥37 wGA is fixed by the constraint that the sum of all birth cohorts must equal 100%.

Data for live births by gestational age are shown in Appendix Table 2. Due to lack of data, births by critical CHD (CCHD) and CLD were based on data from the US as a proxy (Appendix Table 3).

*Appendix Table 2: Live births by gestational age for England and Wales*

| Gestational age at birth (weeks) | | Live births (%) | |
|---|---|---|---|
| Full term | >41 | 1.85% | 92.42% |
| | 41 | 12.79% | |
| | 40 | 23.31% | |
| | 39 | 30.10% | |
| | 38 | 15.57% | |
| | 37 | 8.79% | |
| Late pre-term | 36 | 3.10% | 5.42% |
| | 35 | 1.38% | |
| | 34 | 0.94% | |
| Moderate pre-term | 33 | 0.54% | 0.95% |
| | 32 | 0.40% | |
| Very pre-term | 31 | 0.27% | 0.62% |
| | 30 | 0.21% | |
| | 29 | 0.15% | |
| Extremely pre-term | 28 | 0.13% | 0.59% |
| | <28 | 0.46% | |
| **Source** | **N/A** | **[2] Calculated** | |

*Appendix Table 3: Births by CCHD and CLD for California, United States (2007-2012)*

| Gestational age at birth (weeks) | CCHD births (N) | Non-CCHD births (N) | Fraction of non-CCHD births with CLD (%) | Fraction of births with CCHD or CLD (%) |
|---|---|---|---|---|

| | | | | |
|---|---|---|---|---|
| 32-34 | 343 | 60,872 | 0.40% | 0.96% |
| 29-31 | 163 | 17,951 | 4.40% | 5.26% |
| <29 | 128 | 16,015 | 25.40% | 25.99% |
| **Source** | [3] | | | **Calculated** |

CCHD – critical congenital heart disease; CLD – chronic lung disease

A sample calculation for the final column in Appendix Table 3 is below, i.e., the fraction of 32-34 wGA infants with either CCHD or CLD is

$$\underbrace{\frac{343}{60{,}872 + 343}}_{\text{CCHD}} + \underbrace{\frac{60{,}872}{60{,}872 + 343} \times \frac{0.40}{100}}_{\text{CLD only (no CCHD)}} = 0.96\% \,.$$

The model inputs summarized in Appendix Table 1 can be computed from Appendix Table 2 and Appendix Table 3, see [1, 4].

*Appendix Table 4: Birth cohort by gestational age and high-risk condition*

| Gestational age at birth (weeks) | Infants born ≥34 wGA (%) | Infants born <34 wGA | | Model input |
|---|---|---|---|---|
| | | Without CHD or CLD (%) | With CHD or CLD (%) | |
| ≥37 | 92.42% | N/A | N/A | 92.42% |
| 34-36 | 5.42% | N/A | N/A | 5.42% |
| 32-33 | N/A | 0.94% | 0.01% | 0.94% |
| 29-31 | N/A | 0.59% | 0.03% | 0.59% |
| <29 | N/A | 0.44% | 0.15% | 0.44% |
| High-risk | N/A | N/A | 0.20% | 0.20% |
| **Source** | **[2]** | **Calculated** | | **[2], calculated** |

CHD – congenital heart disease; CLD – chronic lung disease; wGA – weeks gestational age

#### 2. Relative RSV Incidence

The proportion of RSV-positive samples by month, averaged across the 2019/20, 2023/24, and 2024/25 RSV seasons was estimated from the UK Health Security Agency's Respiratory DataMart system [5, 6]. The relative incidence for month X was estimated by computing the ratio of positive samples in month X to positive samples in November, see Appendix Table 5. These input parameters are excluded from the sensitivity analysis.

*Appendix Table 5: Relative RSV incidence*

| **Month** | **Average positive samples (%)** | **Relative incidence** |
|---|---|---|
| January | 6.1% | 0.49 |
| February | 1.9% | 0.15 |
| March | 0.8% | 0.06 |
| April | 0.3% | 0.03 |
| May | 0.4% | 0.03 |
| June | 0.2% | 0.02 |
| July | 0.8% | 0.06 |
| August | 0.9% | 0.08 |
| September | 2.0% | 0.16 |
| October | 7.1% | 0.56 |
| November | 12.6% | 1.00 (reference) |
| December | 9.4% | 0.75 |
| **Source** | **[5-7]** | **Calculation** |

### 3. RSV-H Incidence

RSV-H incidence inputs were stratified by chronological and gestational age and are summarized in Appendix Table 6. Parameters in this table were included in the sensitivity analysis; these parameters were assumed to be Beta distributed with mean given by the value in Appendix Table 6 and with standard deviation assumed to be 10% of the mean.

*Appendix Table 6: RSV-H incidence by chronological and gestational age*

| Chronological age (months) | Gestational age/high-risk condition | | | | | |
|---|---|---|---|---|---|---|
| | High-risk | <29 wGA | 29-31 wGA | 32-34 wGA | 35-36 wGA | ≥37 wGA |
| **RSV-H Incidence** | | | | | | |
| <1 | | 14.47% | 8.83% | 6.37% | 4.93% | 2.78% |
| 2 | | 33.53% | 20.45% | 14.76% | 11.41% | 6.44% |
| 3 | | 25.65% | 15.65% | 11.29% | 8.73% | 4.92% |
| 4 | | 19.06% | 11.63% | 8.39% | 6.49% | 3.66% |
| 5 | | 16.40% | 10.01% | 7.22% | 5.58% | 3.15% |
| 6 | 4.41% | 13.35% | 8.14% | 5.88% | 4.54% | 2.56% |
| 7 | | 10.94% | 6.67% | 4.81% | 3.72% | 2.10% |
| 8 | | 9.01% | 5.49% | 3.96% | 3.06% | 1.73% |
| 9 | | 7.48% | 4.56% | 3.29% | 2.54% | 1.44% |
| 10 | | 6.27% | 3.83% | 2.76% | 2.13% | 1.20% |
| 11 | | 4.74% | 2.89% | 2.09% | 1.61% | 0.91% |
| **RSV-H cases admitted to ICU** | | | | | | |
| <12 | 8.96% | 8.96% | 1.76% | | | |

GA – gestational age; ICU – intensive care unit; RSV-H – respiratory syncytial virus associated hospitalization; wGA – weeks gestational age

#### 3.1. RSV-H for High-Risk Infants

RSV-H incidence for high-risk and non-high-risk infants was estimated by digitizing Supplemental Figure 10 of [5] (estimated from data provided in [8]), see Appendix Table 7. Our model does not stratify high-risk infants by gestational or chronological age, so RSV-H incidence for high-risk infants is a weighted average of the values in Appendix Table 7. Five of the 12 cohorts of <1-month-olds are within the 5-month RSV season, so the weight used for <1-month-olds is 5. Analogously, the weight for 1-7-month-olds is also 5. Only 4 of the 12 cohorts of 8-month-olds are within the 5-month RSV season, thus the weight for 8-month-olds is 4. Analogously, the weights for 9-, 10-, and 11-month-olds are 3, 2, and 1. The weighted average of RSV-H incidence for high-risk infants with the above weights is 4.41%.

*Appendix Table 7: RSV-H incidence by chronological age and risk condition*

| Chronological age (months) | Weights for computing average | Hospitalization incidence | |
|---|---|---|---|
| | | Normal risk | High risk |
| 0 | 5 | 3.01% | 4.06% |
| 1 | 5 | 6.97% | 9.62% |
| 2 | 5 | 5.33% | 7.39% |
| 3 | 5 | 3.96% | 5.38% |
| 4 | 5 | 3.41% | 4.65% |
| 5 | 5 | 2.78% | 3.86% |
| 6 | 5 | 2.27% | 1.84% |
| 7 | 5 | 1.87% | 3.16% |
| 8 | 4 | 1.56% | 2.62% |
| 9 | 3 | 1.30% | 2.12% |
| 10 | 2 | 0.99% | 1.36% |
| 11 | 1 | 0.85% | 1.15% |
| **Average** | **N/A** | **3.41%** | **4.41%** |
| **Source** | **Assumption** | **[5, 8], calculated** | |

#### 3.2. RSV-H Incidence for Non-High-Risk Infants

Coathup and colleagues [9] reported hospitalization incidence (all-cause) by gestational age, see Appendix Table 8. Relative hospitalizations by GA, were computed from the weighted averages. For each chronological age group (<1 month, 1 month, …, 11 months), RSV-H by gestational age was estimated by assuming it was proportional the relative incidence (Appendix Table 8, rightmost column) with proportionality constant chosen to preserve the chronological-age-specific RSV-H incidence (Appendix Table 7).

*Appendix Table 8: All-cause hospitalization by gestational age*

<table>
<tr><th>GA (weeks)</th><th>Hospitalization incidence (%)</th><th>Fraction of birth cohort (%)</th><th>Hospitalization incidence (weighted average; %)</th><th>Relative incidence</th></tr>
<tr><td><28</td><td>6.34%</td><td>0.46%</td><td rowspan="2">5.88%</td><td rowspan="2">5.21</td></tr>
<tr><td>28</td><td rowspan="2">4.26%</td><td>0.13%</td></tr>
<tr><td>29</td><td>0.15%</td><td rowspan="2">3.59%</td><td rowspan="2">3.18</td></tr>
<tr><td>30-31</td><td>3.37%</td><td>0.47%</td></tr>
<tr><td>32</td><td>2.84%</td><td>0.40%</td><td rowspan="2">2.59%</td><td rowspan="2">2.29</td></tr>
<tr><td>33</td><td>2.40%</td><td>0.54%</td></tr>
<tr><td>34</td><td>2.30%</td><td>0.94%</td><td rowspan="3">2.00%</td><td rowspan="3">1.77</td></tr>
<tr><td>35</td><td>1.98%</td><td>1.38%</td></tr>
<tr><td>36</td><td>1.92%</td><td>3.10%</td></tr>
<tr><td>37</td><td>1.63%</td><td>8.79%</td><td rowspan="6">1.13%</td><td rowspan="6">1 (reference)</td></tr>
<tr><td>38</td><td>1.29%</td><td>15.57%</td></tr>
<tr><td>39</td><td>1.10%</td><td>30.10%</td></tr>
<tr><td>40</td><td>1.00%</td><td>23.31%</td></tr>
<tr><td>41</td><td>0.92%</td><td>12.79%</td></tr>
<tr><td>>41</td><td>0.92%</td><td>1.85%</td></tr>
<tr><td>Source</td><td>[9]</td><td>[10]</td><td colspan="2">Calculation</td></tr>
</table>

#### 4. Maternal Vaccine Efficacy

The efficacy for RSVpreF vaccine assumes successful transfer of maternal antibodies from mother to newborn. However, the probability of successful transfer of maternal antibodies depends on both GA at immunization and GA at birth. To estimate the reduction in efficacy as a function of GA the assumptions of Rainisch and colleagues were adopted [11]. Inputs such as marginal distributions for each GA at immunization and GA at were used to estimate the joint distribution for GA at immunization and GA at birth by applying a least squares optimization and the probability of successful transfer of maternal antibodies for GA at birth and for high-risk infants [11, 12].

### 5. RSV-H and RSV-ICU treatment costs

#### 5.1. RSV-H treatment costs

RSV-H cost inputs were stratified by chronological and gestational age and are summarized in Appendix Table 9. Parameters in this table were included in the sensitivity analysis; these parameters were assumed to be Gamma distributed with mean given by the value in Appendix Table 9 and with standard deviation assumed to be 10% of the mean.

*Appendix Table 9: Hospitalization costs by gestational and chronological age*

| Chronological age (months) | Gestational age (weeks) | | | | |
|---|---|---|---|---|---|
| | ≥37 | 34-36 | 32-33 | 29-31 | <29 |
| 0 | £3,489 | £5,505 | £8,687 | £13,709 | £21,633 |
| 1 | £1,791 | £2,825 | £4,459 | £7,036 | £11,103 |
| 2 | £1,610 | £2,541 | £4,010 | £6,327 | £9,984 |
| 3 | £1,452 | £2,291 | £3,616 | £5,706 | £9,004 |
| 4 | £1,403 | £2,214 | £3,493 | £5,512 | £8,698 |
| 5 | £1,267 | £1,999 | £3,154 | £4,978 | £7,855 |
| 6 | £1,217 | £1,921 | £3,032 | £4,784 | £7,549 |
| 7 | £1,363 | £2,150 | £3,393 | £5,354 | £8,449 |
| 8 | £1,324 | £2,089 | £3,296 | £5,202 | £8,208 |
| 9 | £1,339 | £2,113 | £3,335 | £5,263 | £8,304 |
| 10 | £1,526 | £2,408 | £3,800 | £5,996 | £9,462 |
| 11 | £1,245 | £1,964 | £3,099 | £4,891 | £7,718 |

We estimated the cost per hospitalized day for RSV-H from NHS Reference costs. Specifically, the hospitalization costs for Paediatric Acute Bronchiolitis (code: PD15) was estimated to be £1,881.14 by taking a weighted average of codes PD15A, PD15B, PD15C, and PD15D. Assuming 2.34 hospital bed days per infection, the cost per hospitalized day for RSV-H was £1,881.14 / 2.34 = £803.91.

Total RSV-H costs were estimated by multiplying the number of days hospitalized by the daily cost of hospitalization, i.e., £803.91. The number of hospital bed days was estimated from results published by Reeves and colleagues [8], see Appendix Table 10. For example, we computed the average number of bed days per hospital admission for normal-risk infants using the formula

$$\frac{\#\ bed\ days \times 79\%}{\#\ admissions \times 95\%}$$

and proceed analogously for high-risk infants. Results are summarized in Appendix Table 11.

*Appendix Table 10: Annual hospitalization admissions and bed days [8]*

| Parameter | Admissions | | Bed days | |
|---|---|---|---|---|
| | n | (%) | n | (%) |
| **Clinical risk group** | | | | |

| | | | | |
|---|---|---|---|---|
| No | 19,415 | 95% | 45,747 | 79% |
| Yes | 944 | 5% | 12,160 | 21% |
| **Chronological age (months)** | | | | |
| <1 | 1,772 | 8.70% | 10,529 | 18.18% |
| 1 | 4,174 | 20.50% | 12,729 | 21.98% |
| 2 | 3,184 | 15.64% | 8,729 | 15.07% |
| 3 | 2,323 | 11.41% | 5,745 | 9.92% |
| 4 | 2,013 | 9.89% | 4,809 | 8.30% |
| 5 | 1,661 | 8.16% | 3,584 | 6.19% |
| 6 | 1,359 | 6.67% | 2,819 | 4.87% |
| 7 | 1,121 | 5.51% | 2,601 | 4.49% |
| 8 | 911 | 4.47% | 2,053 | 3.55% |
| 9 | 771 | 3.79% | 1,759 | 3.04% |
| 10 | 580 | 2.85% | 1,508 | 2.60% |
| 11 | 493 | 2.42% | 1,045 | 1.80% |

*Appendix Table 11: RSV non-ICU hospitalization costs by chronological age*

| Chronological age (months) | Hospital bed days | | Hospitalization cost | | |
|---|---|---|---|---|---|
| | Normal risk | High risk | Normal risk | High risk | Average |
| <1 | 4.92 | 26.91 | £3,955 | £21,633 | £4,775 |
| 1 | 2.53 | 13.81 | £2,034 | £11,102 | £2,454 |
| 2 | 2.27 | 12.42 | £1,825 | £9,985 | £2,203 |
| 3 | 2.05 | 11.20 | £1,648 | £9,004 | £1,989 |
| 4 | 1.98 | 10.82 | £1,592 | £8,698 | £1,921 |
| 5 | 1.79 | 9.77 | £1,439 | £7,854 | £1,736 |
| 6 | 1.72 | 9.39 | £1,383 | £7,549 | £1,669 |
| 7 | 1.92 | 10.51 | £1,544 | £8,449 | £1,864 |
| 8 | 1.87 | 10.21 | £1,503 | £8,207 | £1,814 |
| 9 | 1.89 | 10.33 | £1,519 | £8,304 | £1,834 |
| 10 | 2.15 | 11.77 | £1,728 | £9,462 | £2,087 |
| 11 | 1.76 | 9.60 | £1,415 | £7,718 | £1,707 |

To estimate hospitalizations by chronological and gestational age we assume that

(a) hospitalization costs for <29 wGA infants are equal to hospitalization costs of high-risk infants,
(b) average hospitalization costs for ≥29 wGA infants is equal to hospitalization costs for normal-risk infants, and
(c) there are parameters $\{cost^{a}_{\geq 37}\}^{11}_{a=0}$ and $m$ such that the hospitalization cost for an infant with gestational age g and chronological age a is

$$cost^{a}_{\geq 37} \times m^{g-1} \,,$$

where $g = 1, 2, 3, 4, 5$ for gestational age groups ≥37 wGA, 34-36 wGA, 32-33 wGA, 29-31 wGA, and <29 wGA, respectively.

Under these assumptions the hospitalization cost for high-risk infants aged $a$ months is

$$cost^{a}_{\geq 37} \times m^{4} .$$

The hospitalization cost for normal-risk infants aged $a$ months is the weighted average

$$\sum_{g=1}^{4} w_g \times costs^{a}_{\geq 37} \times m^{g-1} ,$$

where weights are given below in Appendix Table 12.

*Appendix Table 12: Weights for computing hospitalization costs by gestational age*

| Gestational age (weeks) | Relative hospitalization incidence | Population (%) | Hospitalizations (rel. hosp. incidence x population) | Weights ($w_g$; proportional to hospitalizations) |
|---|---|---|---|---|
| ≥ 37 | 1 | 92.42% | 0.9242 | 0.8715 |
| 34-36 | 1.77 | 5.42% | 0.0959 | 0.0905 |
| 32-33 | 2.29 | 0.94% | 0.0215 | 0.0203 |
| 29-31 | 3.18 | 0.59% | 0.0188 | 0.0177 |
| **Source** | **Appendix Table 8** | | **Calculated** | **Calculated** |

We choose the parameters $\{cost^{a}_{\geq 37}\}_{a=0}^{11}$ and $m$ to minimize the sum of squared differences between hospitalization costs for normal-risk infants and high-risk infants, i.e., we minimize the objective function

$$\sum_{a=0}^{11} \left(costs^{a}_{\geq 37} \times m^{4} - \text{high-risk hospitalization cost for age } a\right)^2$$
$$+ \sum_{a=0}^{11} \left( \sum_{g=1}^{4} w_g \times costs^{a}_{\geq 37} \times m^{g-1} - \text{normal-risk hospitalization cost for age } a \right)^2 .$$

Results from the calibration are recorded in Appendix Table 13 and Appendix Table 9. Appendix Figure 1 plots hospitalization costs from Appendix Table 11 versus calibrated estimates.

*Appendix Table 13: Calibrated parameters for hospitalization costs by chronological and gestational age*

| Parameter | Value |
|---|---|
| $m$ | 1.578 |
| $costs^{0}_{\geq 37}$ | £3,489 |
| $costs^{1}_{\geq 37}$ | £1,791 |
| $costs^{2}_{\geq 37}$ | £1,610 |
| $costs^{3}_{\geq 37}$ | £1,452 |
| $costs^{4}_{\geq 37}$ | £1,403 |
| $costs^{5}_{\geq 37}$ | £1,267 |
| $costs^{6}_{\geq 37}$ | £1,217 |

| | |
|---|---|
| $costs^{7}_{\geq 37}$ | £1,363 |
| $costs^{8}_{\geq 37}$ | £1,324 |
| $costs^{9}_{\geq 37}$ | £1,339 |
| $costs^{10}_{\geq 37}$ | £1,526 |
| $costs^{11}_{\geq 37}$ | £1,245 |

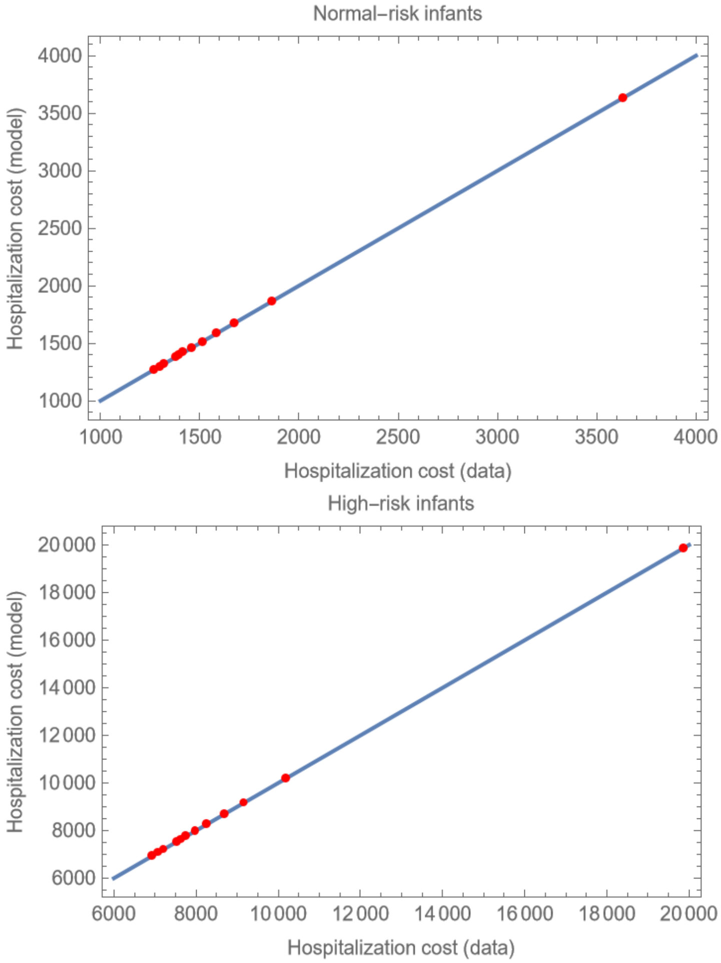

*Appendix Figure 1: Predicted versus observed hospitalization costs for normal- and high-risk infants*

#### 5.2. RSV-ICU treatment costs

Marginal costs for RSV-ICU (i.e., the difference between RSV-H and RSV-ICU costs) was estimated to be £5,282. We estimate the average non-ICU hospitalization cost from Appendix Table 9 by applying the following weights, see Appendix Table 14. There are 5 monthly birth cohorts that are <1 months during their first RSV season. Similarly, there are 5 monthly birth cohorts that are 1-, 2-, 3-, 4-, 5-, 6-, and 7-months during their first RSV season. In contrast, there are 4, 3, 2, and 1 birth cohorts that are 8-, 9-, 10-, and 11-months during their first RSV season. RSV ICU costs comprise average hospitalization costs plus the marginal ICU admission cost, see Appendix Table 15.

*Appendix Table 14: Weights used for computing average of RSV-H (non-ICU) costs*

| Chronological age (months) | Weight | Weight (normalized) |
|---|---|---|
| <1 | 5 | 0.1 |
| 1 | 5 | 0.1 |
| 2 | 5 | 0.1 |
| 3 | 5 | 0.1 |
| 4 | 5 | 0.1 |
| 5 | 5 | 0.1 |
| 6 | 5 | 0.1 |
| 7 | 5 | 0.1 |
| 8 | 4 | 0.08 |
| 9 | 3 | 0.06 |
| 10 | 2 | 0.04 |
| 11 | 1 | 0.02 |

*Appendix Table 15: RSV-ICU treatment costs by gestational age*

| Gestational age (weeks) | RSV ICU cost |
|---|---|
| **≥37** | £6,913 |
| **34-36** | £7,856 |
| **32-33** | £9,344 |
| **29-31** | £11,692 |
| **<29** | £15,397 |
| **Source** | [13] |

### 6. RSV-RW costs

Shefali-Patel and colleagues [14] reported excess costs of £1,342 GBP over a 2-year follow-up period for 33-35 wGA infants infected with RSV. We inflate this value to 2024 GBP [15] and halved it, giving a 1-year follow up figure of £1,052 for infants born 29-35 wGA. For infants born <29 wGA and ≥37 wGA we use a multiplier by comparing to Chirikov and colleagues [16], see Appendix Table 16.

*Appendix Table 16: Excess RSV-RW costs (1-year follow-up)*

| Gestational age (weeks) | RSV-RW costs (1-year follow-up from index RSV case) |
|---|---|
| ≥37 | £678 |
| 35-36 | £1,052 |
| 32-34 | |
| 29-31 | |
| <29 | £2,603 |
| **Source** | **[14, 16], calculated** |

Table values rounded up to nearest whole 2024 GBP.

### 7. Additional results

Under seasonal maternal vaccination strategies pregnant individuals whose due dates occurred during the RSV season were eligible for vaccination. Model epidemiological and cost results for in-season administration of maternal vaccine are shown in Appendix Table 17 and Appendix Table 18, respectively. Deterministic sensitivity analyses results for incremental QALYs and incremental treatment costs for seasonal plus catch-up administration of clesrovimab versus in-season maternal vaccination are shown in Supplement Figure 1 and Supplement Figure 2.

*Appendix Table 17. Incremental change in RSV-associated clinical outcomes resulting from seasonal, seasonal plus catch-up, and year-round administration of clesrovimab to all infants, as compared with administration of maternal vaccine to mothers of all infants born in-season.*

| Administration | Clinical outcome | Clesrovimab (75.1% MALRI efficacy) vs |
|---|---|---|
| | | Maternal vaccine* |
| Seasonal | RSV-H-noICU | -2,813 (-16.3%) |
| | RSV-ED | -4,303 (-10.3%) |
| | RSV-GP | -8,221 (-15.6%) |
| | RSV-H-ICU | -56 (-16.7%) |
| | RSV-RW | -596 (-14.4%) |
| | RSV Deaths | -2 (-16.4%) |
| Seasonal plus catch-up | RSV-H-noICU | -9,642 (-56.0%) |
| | RSV-ED | -20,100 (-48.0%) |
| | RSV-GP | -29,383 (-55.8%) |
| | RSV-H-ICU | -187 (-56.1%) |
| | RSV-RW | -2,203 (-53.2%) |
| | RSV Deaths | -6 (-56.0%) |
| Year-round | RSV-H-noICU | -7,172 (-41.6%) |
| | RSV-ED | -14,221 (-33.9%) |
| | RSV-GP | -24,685 (-46.9%) |
| | RSV-H-ICU | -139 (-41.9%) |
| | RSV-RW | -1,706 (-41.2%) |
| | RSV Deaths | -4 (-41.7%) |

ICU – intensive care unit; RSV-ED – RSV-associated ED visits; RSV-H – RSV-associated hospitalizations; RSV-O – RSV-associated outpatient visits; RSV-RW – RSV-associated recurrent wheezing

* Administration of maternal vaccine is seasonal only.

*Appendix Table 18. Incremental change in RSV-associated costs resulting from seasonal, seasonal plus catch-up, and year-round administration of clesrovimab to all infants, as compared with administration of maternal vaccine to mothers of all infants born in-season. (2024 GBP)*

| Administration | Cost outcome | Clesrovimab (75.1% MALRI efficacy) vs |
|---|---|---|
| | | Maternal vaccine* |
| Seasonal | RSV-H-noICU | -£7,675,344 (-22.3%) |
| | RSV-ED | -£1,174,468 (-10.3%) |

| | | |
|---|---|---|
| | RSV-GP | -£386,565 (-15.6%) |
| | RSV-H-ICU | -£451,513 (-17.3%) |
| | RSV-RW | NA |
| | Total treatment cost | -£9,687,890 (-19.0%) |
| | Doses administered | 94,385 |
| | QALY loss averted | 104 (-14.7%) |
| **Seasonal plus catch-up** | RSV-H-noICU | -£19,531,908 (-56.7%) |
| | RSV-ED | -£5,486,665 (-48.0%) |
| | RSV-GP | -£1,381,594 (-55.8%) |
| | RSV-H-ICU | -£1,470,627 (-56.4%) |
| | RSV-RW | NA |
| | Total treatment cost | -£27,870,794 (-54.7%) |
| | Doses administered | 422,367 |
| | QALY loss averted | 381 (-54.0%) |
| **Year-round** | RSV-H-noICU | -£15,294,770 (-44.4%) |
| | RSV-ED | -£3,881,908 (-33.9%) |
| | RSV-GP | -£1,160,677 (-46.9%) |
| | RSV-H-ICU | -£1,100,548 (-42.2%) |
| | RSV-RW | NA |
| | Total treatment cost | -£21,437,902 (-42.1%) |
| | Doses administered | 422,367 |
| | QALY loss averted | 292 (-41.4%) |

ICU – intensive care unit; RSV-ED – RSV-associated ED visits; RSV-H – RSV-associated hospitalizations; RSV-O – RSV-associated outpatient visits; RSV-RW – RSV-associated recurrent wheezing

* Administration of maternal vaccine is seasonal only.

*Supplement Figure 1. Incremental QALYs one-way sensitivity analysis for seasonal with catch-up administration of clesrovimab versus in-season maternal vaccination.*

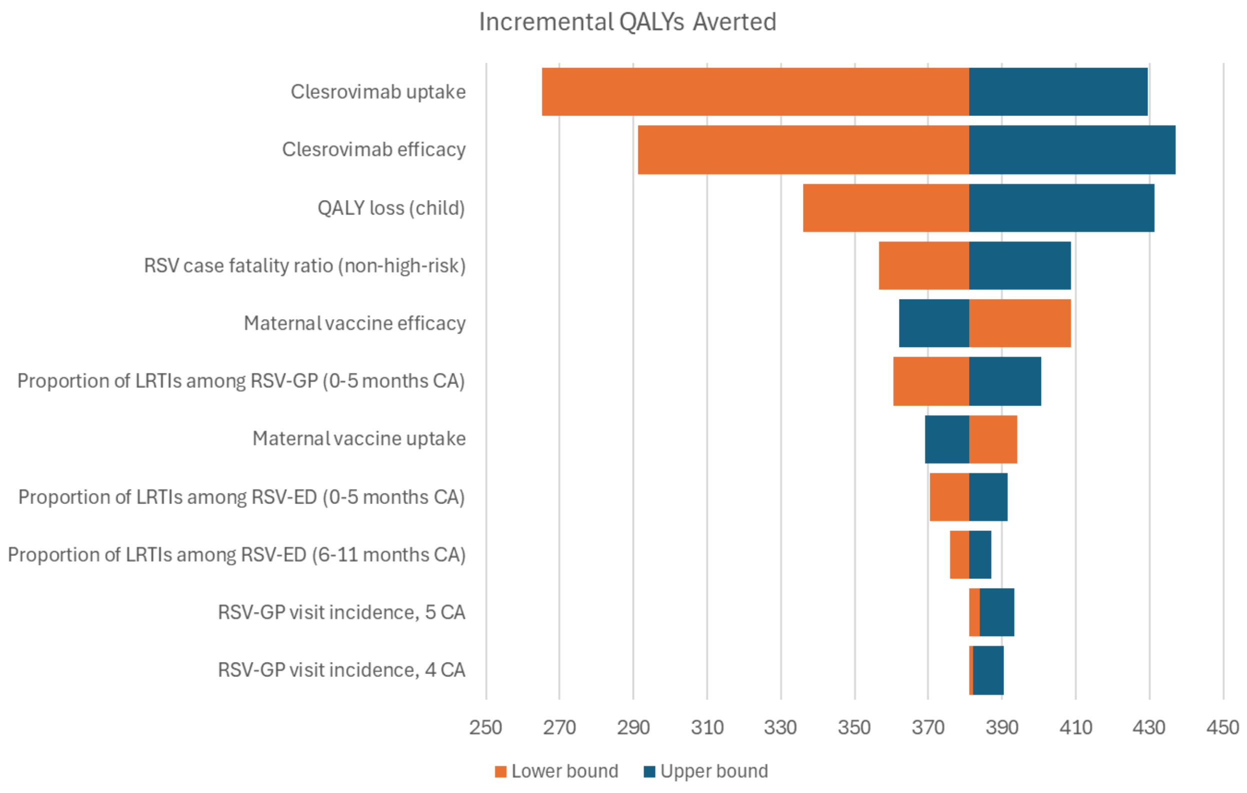

*Supplement Figure 2. Incremental treatment costs one-way sensitivity analysis for seasonal plus catch-up administration of clesrovimab versus in-season maternal vaccination. (2024 GBP)*

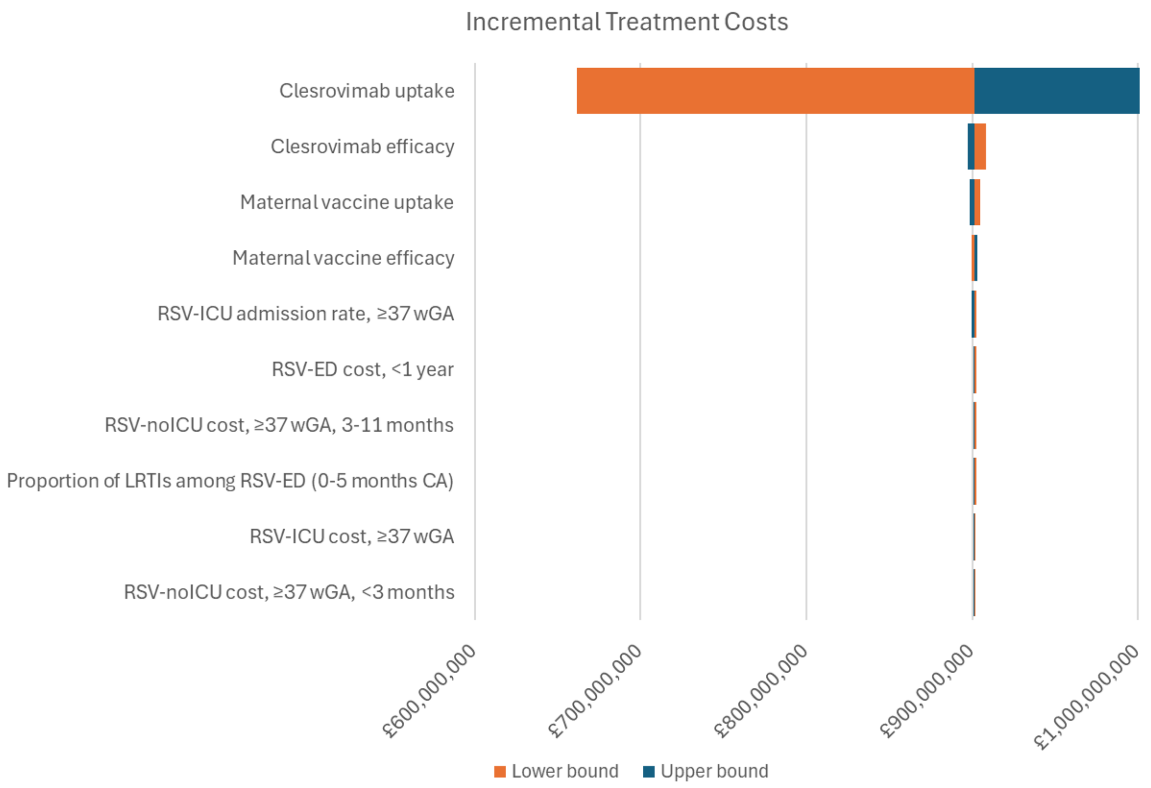
Incremental Treatment Costs
Clesrovimab uptake
Clesrovimab efficacy
Maternal vaccine uptake
Maternal vaccine efficacy
RSV-ICU admission rate, ≥37 wGA
RSV-ED cost, <1 year
RSV-noICU cost, ≥37 wGA, 3-11 months
Proportion of LRTIs among RSV-ED (0-5 months CA)
RSV-ICU cost, ≥37 wGA
RSV-noICU cost, ≥37 wGA, <3 months
£600,000,000
£700,000,000
£800,000,000
£900,000,000
£1,000,000,000
Lower bound
Upper bound
